# Renewable Energy Use in Australian Public Hospitals

**DOI:** 10.1101/2021.02.25.21252432

**Authors:** Hayden Burch, Matthew Anstey, Forbes McGain

## Abstract

**Objective:** Are Australian hospitals moving towards renewable energy sources for their electricity, and aligning energy choices with core business, i.e. protecting and promoting health?

**Design:** Cross-sectional analysis of Australian state/territory amalgamated energy data

**Setting:** Healthcare’s carbon footprint is approximately 7% of Australia’s total carbon footprint. It is unknown if Australian public hospitals are decoupling energy needs from carbon emissions over and above state/territory based renewable energy targets.

**Participants:** 693 Australian public hospitals direct energy usage (renewable & non-renewable electricity [produced/purchased], natural gas, liquefied petroleum gas, for the three consecutive years from 2016/17 to 2018/19.

**Main outcome measures:** All direct energy produced/purchased and consumed (converted to kilowatt-hours).

**Results:** Australian public hospitals consumed 4,122 gigawatt-hours of energy in 2018/19. Electricity use was 2,504 (61%) GWh, natural gas 1,436 (35%) GWh and renewable energy 94 GWh (2.3%). Victoria and New South Wales combined consumed 2,494/ 4,122 GWh (60%) of total Australian public healthcare energy but each produced/purchased less than 1% renewable electricity. For Queensland, a Health GreenPower purchase comprised the majority (71/94 GWh; 76%) of renewable energy production/purchase by all Australian public hospitals. By comparison, individual tertiary education institutions produced/purchased more renewable energy than all Australian public hospitals combined (University of NSW 124 GWh/yr, Swinburne University 90 GWh/yr, 2018/19).

**Conclusions:** Australian public hospitals obtain approximately 2.3% of total energy from renewable electricity. One third of hospital energy use stems from fossil gas use. The Australian public hospital system has no documented plans to transition to renewable energy, contrasting with the University sector.

The known
Australian healthcare contributes approximately 7% of Australia’s total carbon footprint with public hospital energy use a major source of healthcare associated carbon emissions.

The new
Australian public hospitals consumed 4122 gigawatt-hours in 2018/2019. Approximately 2.3% (94/4,122 gigawatt-hours) of hospital energy was sourced from renewables, beyond state-wide renewable electricity penetration.

The implications
Australian public hospitals are large emitters of greenhouse gases. Hospital fossil fuel energy use and subsequent pollution continues unabated. Such increasing pollution is at odds with the ethos ‘first do no harm’.

## Introduction

Increases in extreme weather in Australia are contributing to adverse physical, mental and intergenerational health outcomes (1, 2). Health systems themselves are increasingly at direct risk from the climate crisis (2). Yet healthcare itself pollutes: currently the Australian healthcare system contributes approximately 7% of Australia’s total carbon footprint (3). Direct energy use (coal-generated electricity and natural/fossil gas) are large sources of healthcare CO_2_ equivalent (CO_2_e) emissions (3), with coal-generated electricity additionally contributing to air particulate matter with strong associations with cardiorespiratory illnesses (2). Hospitals in particular are large consumers of energy; with public hospitals representing over half of public-sector energy consumption in most states and territories (4, 5). Hospitals require continuous operation, with large energy demands primarily from heating, ventilation and air conditioning (HVAC) (6).

Since 2018, 21 healthcare institutions in 12 countries have signed on to a 100% renewable electricity target for their healthcare facilities (6). Kaiser Permanente, the United States’ largest non-profit healthcare system, obtains over 1,000 gigawatt-hours (GWh) of *GreenPower* (purchased large-scale renewable electricity) and is carbon net positive (7). The UK’s *National Health Service Sustainable Development Unit* has reduced health sector’s CO_2_e emissions by 11% between 2007-2015, despite an 18% increase in inpatient admissions (8).

Hospitals can address energy use in two ways: by increasing efficiency and by transitioning electricity demand to renewable sources (directly through electricity production from solar, wind, waste fuels, and hydrogen, or indirectly, e.g. purchased *GreenPower)* (4). Co- or tri-generation electricity are energy-efficiency variants, relying on fossil/natural gas. Energy efficiency is highly important and routinely has rapid payback times (5, 9). However, in Victoria, for example, whilst energy efficiency strategies achieved a 9% reduction in CO_2_e emissions/m^2^ of hospital floor-space between 2005-2018, overall energy use grew 22% and carbon emissions by 32% due to rising demand (10).

Since 2017 public hospitals have been included in both the *National Built Environment Rating System* (NABERS) (11) and *National Greenhouse and Energy Reporting* (NGER) schemes (12). Under NABERS, 274 of 693 Australian public hospitals are included in 2018/19 NABERS analysis, though none are publicly disclosed (11). Under NGER, hospitals must report if energy consumption is 100TJ or more per year and/or if emissions equal 25kt CO_2_e emissions/annum (i.e. hospitals with at least 200 acute beds) (12). Despite these reporting mechanisms, it is unclear how rapidly Australian hospitals are moving towards renewable energy alternatives and what progress has been made by hospitals toward the carbon emission aspirations/targets established by each Australian state/territory.

The objective of this study was to evaluate the total energy use, electricity use, fossil gas use, and renewable electricity generation/purchase by Australian public hospitals. We compare the performance of Australian healthcare to international healthcare leaders and the university sector.

## Methods

We examined Australian state/territory public hospital direct energy data for the decade 2010-19. Although we sought 10 years of data, we were only able to obtain complete data for all states/territories for three consecutive years 2016/17 – 2018/19. There were 693 public hospitals in Australia in 2018/19 (Australian Institute of Health and Welfare data) (13).

We sought data from two sources: Australian State/Territory Health Departments, and the Australian national Clean Energy Regulator (responsible for NGER reporting). We wrote to Australian state/territory Health Departments in August-October 2019 seeking annualised data for public hospital direct energy use. We also wrote to three health services in Western Australia (within Eastern-, North- and South-Metropolitan regions) as no renewable electricity data could be identified by the WA Health Department. Hospital-level information was amalgamated into state-based calculations to avoid inadvertent identification of individual hospitals. Tertiary education was selected as a comparator with similarly large multi-building institutions, and considerable energy demands (4).

The following Australian public hospital data was requested for 2010-2019: total energy use, fossil/natural gas use, renewable and non-renewable electricity (produced or purchased, e.g. rooftop solar photovoltaic (PV), GreenPower) in kilowatt-hours (kWh), natural gas (gigajoules), liquefied petroleum gas (LPG) (kilolitres), co/tri-generation (kWh)). We converted all energy data into kWh for simplification, i.e. for fossil/natural gas; 1GJ= 278kWh, for LPG; 1kL= 6,900kWh (14). Solar PV data supplied as kilowatts/kilowatt-peak were multiplied by the hourly daily solar average per state/territory capital city for 365 operating days (15). Peak-watts were used to calculate a best-case scenario. If raw gas co-generation electricity data were not provided, we multiplied total kilowatt-peak capacity by the minimum annual running time to justify operation as per a NSW government approach (3,300 hours p.a. or 38% total time usage) (16). Solar hot water production was included in Queensland supplied data, and approximated by researchers in South Australia as 2kWh/panel/day (4) and not reported elsewhere.

In order to establish the active contribution of the healthcare industry to reducing fossil fuel use, we focussed upon renewable electricity generated/purchased by the healthcare sector. Only health sector purchased GreenPower was attributed to hospital renewable energy calculations. That is, we only included production/purchase of renewable electricity beyond the state’s/territory’s grid electricity. We did not include diesel fuel use (used either for transport, back-up power or in smaller rural hospitals for electricity generation) due to poor data access.

## Results

We received responses from all state/territory Health Departments and from metropolitan Western Australian health services from November 2019 to February 2020. Data were robust for 2016/17 to 2018/19. Data prior to 2016/17 were: unavailable, unreliable or not inclusive of all hospitals. We also received data supplied under NGER (the Clean Energy Regulator), but such data were unreliable (high reporting thresholds and heavily redacted due to concerns about revealing individual hospitals).

Total Australian hospital energy use was stable for the three years (2016/17 to 2018/19 (Table 1). From 2016/17 to 2018/19 Australian public hospitals increased production/purchase of renewable energy from 14/ 4,132 (0.3%) GWh to 94/ 4,122 (2.3%) GWh (Table 1). Australian renewable grid electricity uptake grew by 8.3% (from 15.7% in 2016/17 to 24.0% in 2018/19).

**Table 1.**
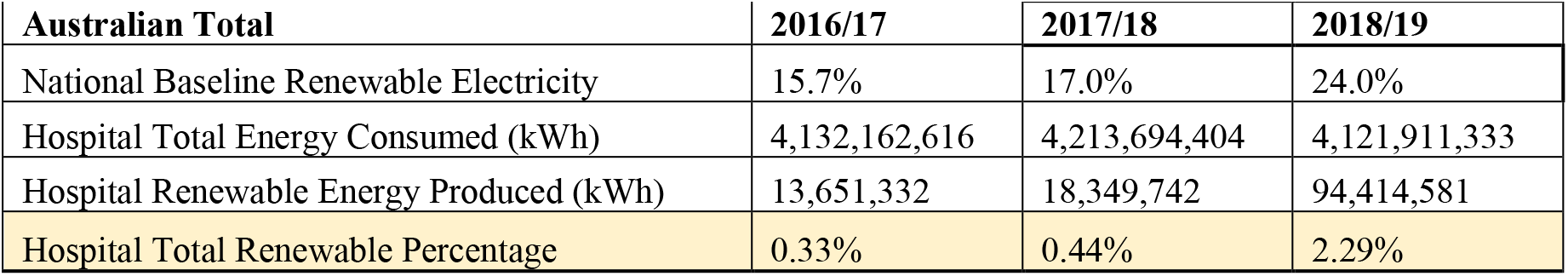
**Australian national grid renewable electricity baseline, public hospitals’ total direct energy consumed, renewable energy produced, and percentage renewable, 2016/17 to 2018/19**

On a national level, grid electricity usage was the majority of total energy consumed (range 2,495-2,507 [59.2-60.7% of total energy] GWh), incorporating GreenPower certificates (range 1.01-72.69 [0.02-1.8%] GWh). Natural gas use was also large (range 1,444-1,522 [34.9-36.1%] GWh), followed by liquid propane gas (range 95-107 [2.3-2.6%] GWh), co/tri-generation (range 62.0-73.6 [1.5-2.1%] GWh), and rooftop solar PV (range 12.6-20.9 [0.3-0.5%] GWh). No biofuels were used according to discussions with state health representatives.

State/Territory public hospital total energy, renewable electricity use and grid renewable electricity baseline is shown (Table 2). A year-on-year comparison of total energy consumption varies for each state/territory over the three-year period. In Victoria, the decrease in total energy consumed between 2017/18 and 2018/19 can be attributed to a reduction in natural gas, LPG and grid electricity usage (potentially due to improved hospital energy efficiency).

**Table 2.** **State baseline grid renewable electricity baseline, public hospital total energy consumed, renewable energy produced, and percentage, by state/territory 2016/17 to 2018/19**

| State | Energy (kWh) | 2016/17 | 2017/18 | 2018/19 |
| --- | --- | --- | --- | --- |
| VIC | State baseline renewables | 12.0% | 15.2% | 23.9% |
|  | Total hospital energy consumed | 1,305,587,305 | 1,310,007,973 | 1,288,291,614 |
|  | Hospital renewable energy produced | 3,287,858 | 7,679,177 | 11,056,842 |
|  | Hospital percentage renewable | 0.3% | 0.6% | 0.9% |
| NSW | State baseline renewables | 16.0% | 15.7% | 17.1% |
|  | Total hospital energy consumed | 1,194,656,992 | 1,218,508,959 | 1,206,019,026 |
|  | Hospital renewable energy produced | 7,053,271 | 7,035,874 | 7,007,253 |
|  | Hospital percentage renewable | 0.6% | 0.6% | 0.6% |
| QLD | State baseline renewables | 6.5% | 7.2% | 14.1% |
|  | Total hospital energy consumed | 773,110,761 | 770,316,605 | 778,594,708 |
|  | Hospital renewable energy produced | 2,136,352 | 2,121,083 | 73,694,459 |
|  | Hospital percentage renewable | 0.3% | 0.3% | 9.5% |
| WA | State baseline renewables | 7.5% | 7.7% | 20.9% |
|  | Total hospital energy consumed | 336,494,022 | 354,697,727 | 335,783,040 |
|  | Hospital renewable energy produced | 0 | 0 | 395,558 |
|  | Hospital percentage renewable | 0.0% | 0.0% | 0.1% |
| ACT | State baseline renewables | 18.1% | 28.2% | 43.9% |
|  | Total hospital energy consumed | 70,195,556 | 70,310,833 | 67,426,111 |
|  | Hospital renewable energy produced | 549,140 | 828,837 | 811,332 |
|  | Hospital percentage renewable | 0.8% | 1.2% | 1.2% |
| TAS | State baseline renewables | 91.3% | 91.3% | 95.6% |
|  | Total hospital energy consumed | 71,838,719 | 70,987,458 | 71,395,800 |
|  | Hospital renewable energy produced | 414,690 | 428,760 | 299,971 |
|  | Hospital percentage renewable | 0.6% | 0.6% | 0.4% |
| NT | State baseline renewables | 2.9% | 3.5% | 6.0% |
|  | Total hospital energy consumed | 54,281,807 | 56,414,704 | 49,612,525 |
|  | Hospital renewable energy produced | - | - | - |
|  | Hospital percentage renewable | 0.0% | 0.0% | 0.0% |
| SA | State baseline renewables | 47.0% | 46.6% | 52.1% |
|  | Total hospital energy consumed | 325,997,454 | 362,450,144 | 324,788,510 |
|  | Hospital renewable energy produced | 210,021 | 256,011 | 1,149,166 |
|  | Hospital percentage renewable | 0.1% | 0.1% | 0.4% |

Blue - data sourced directly from Western Australian health services.

Victoria and New South Wales combined consumed 60% of total Australian public hospital energy (1,288 [31.3%] and 1,206 [29.3%] of 4,122 GWh respectively, 2018/19) (Figure 1). Queensland public hospitals consumed the third largest amount of energy (778/4,122 [18.9%] GWh) and produced/purchased the most renewable electricity (73.7/778 GWh [9.5%] of state public hospital energy) in 2018/19.

**Figure 1.**
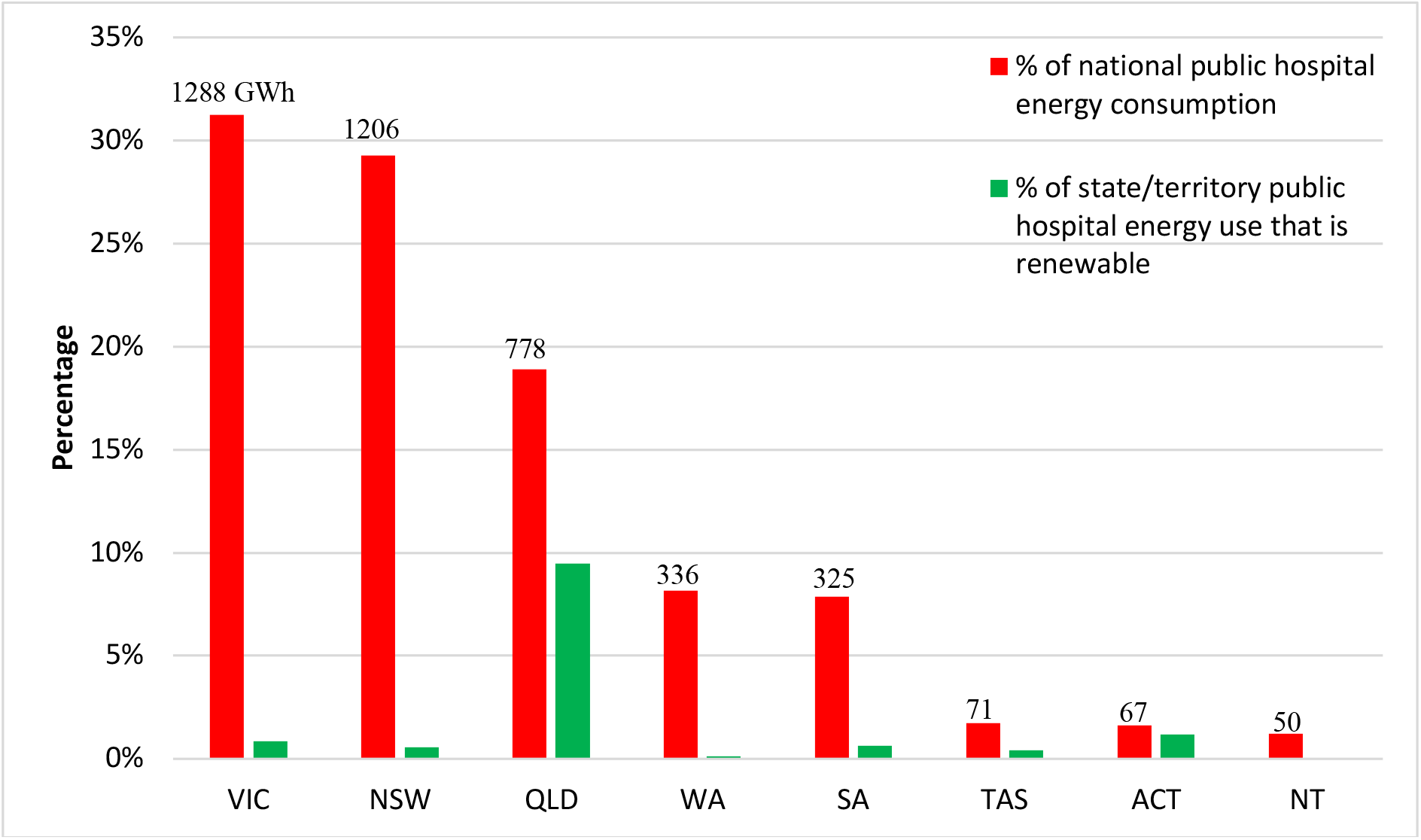
**Percentage consumption of national public hospital energy by each state/territory, and percentage state/territory level production of renewable energy, 2018/19**

Figure 2 provides visual representation of the percentage uptake of renewables by state/territory public hospitals. The large increase in renewable energy in 2018/19 was a result of Queensland Health being a benefactor of a whole-of-government GreenPower Purchase Agreement (71.4 GWh purchased by Queensland Health). Aside from Queensland Health, renewable energy uptake across remaining states/territories is relatively small to non-existent (range 0 – 1.2%, 2018-19).

**Figure 2.**
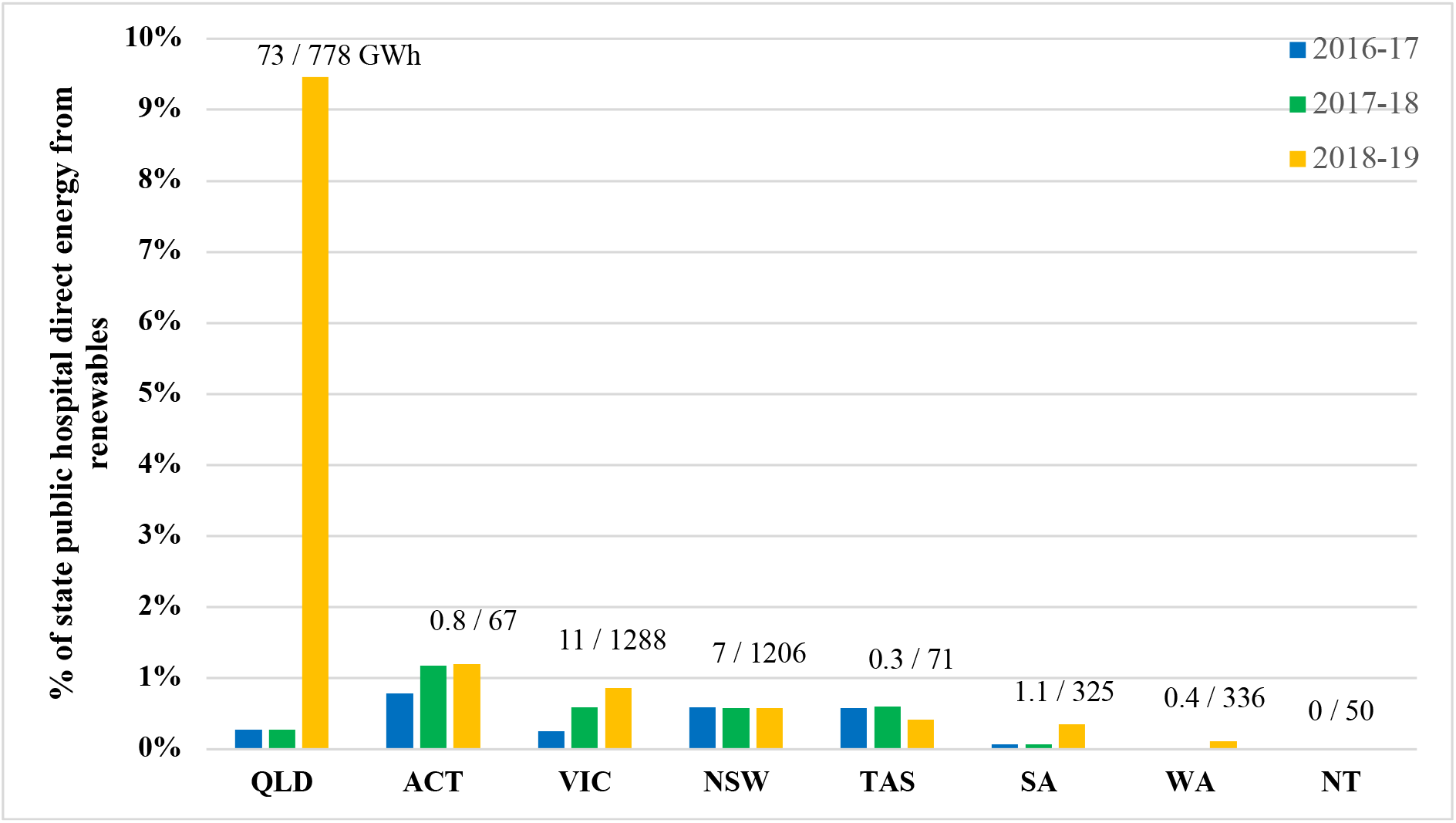
Percentage of state/territory public hospital direct energy use from renewable sources over and above state-baseline renewable penetration (2016/17 to 2018/19) *Large increase in Qld due to whole-of-government purchase of GreenPower.

Victorian public hospitals increased renewable electricity from 0.3% to 0.9% (3.3-11.1 GWh/yr) between 2016/17 to 2018/19. This 7.8 GWh increase by 2018/19 above 2016/17 uptake was accompanied by a 17.3 GWh (1.3%) increase in energy efficiency over the same period. In 2018/19 New South Wales public hospitals produced 7.0/1,206 (0.58%) GWh of renewable energy, unchanged compared to 7.01 (0.59%) GWh in 2016/17, whilst total NSW hospital energy consumption increased by 11.4 GWh.

Figure 3 compares of state/territory grid electricity versus natural gas usage. In Victoria, South Australia and Australia Capital Territory approximately half of total energy consumption is by natural/fossil gas.

**Figure 3.**
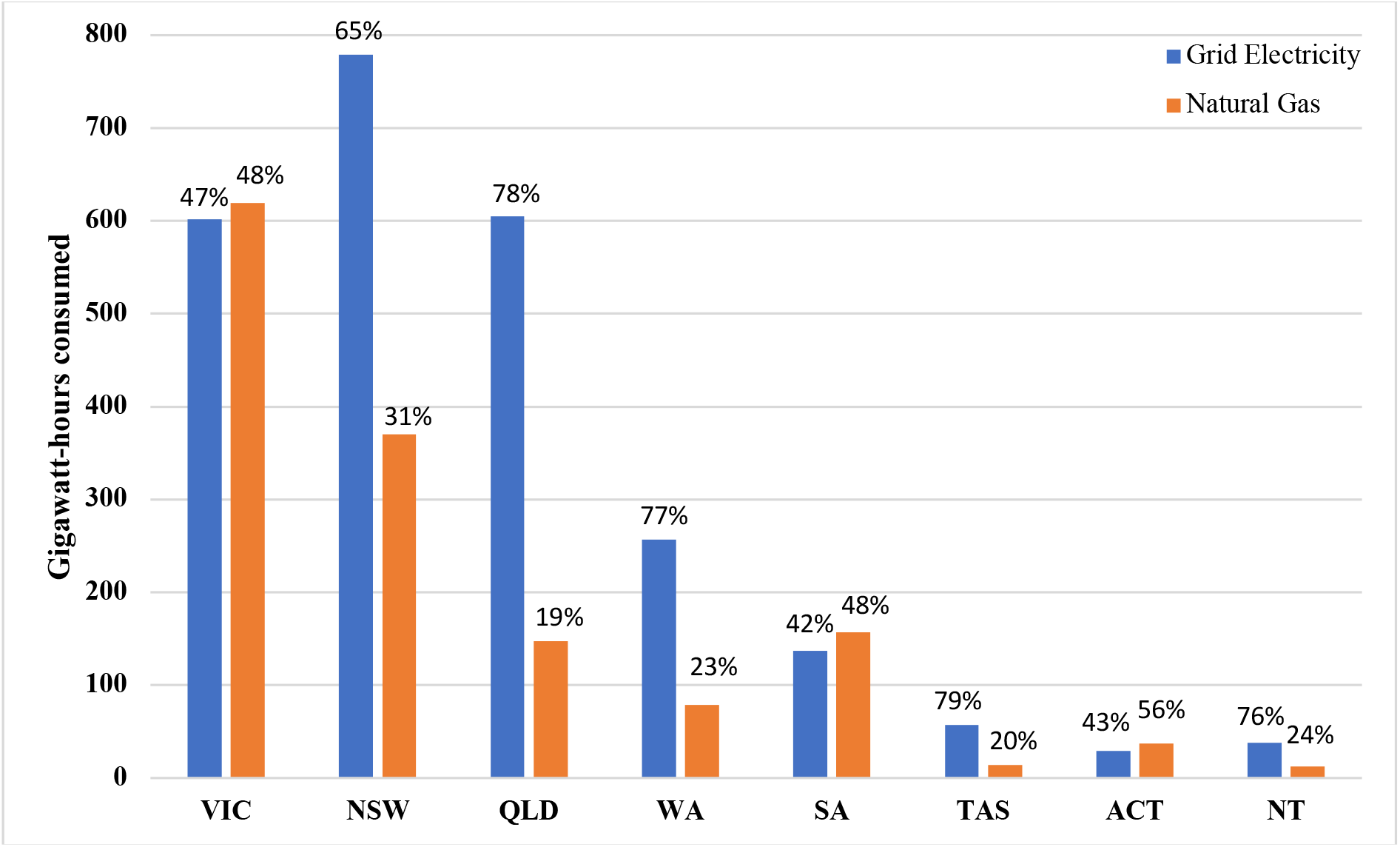
Public hospital energy consumption derived from grid electricity versus natural gas, per state/territory, 2018/19. *Grid electricity includes baseline grid electricity and purchased hospital GreenPower. Percentages may not sum to 100% as not all forms of energy are included (e.g. LPG, co-generation electricity, solar hot water).

In recognising the 2016 Paris Climate Accord, Australia’s states and territories have established net zero emissions and renewable energy targets (Table 3) (17). In some states/territories additional health sector targets are included (10, 18-22).

**Table 3.** **National, State and Territory Renewable Electricity Generation, Renewable Electricity, and Net Zero CO2e Emissions Targets, and Health Sector specific ambitions**

| Jurisdiction | % Grid renewable electricity (2019) | 2030 target | Net zero emissions target | Health sector targets |
| --- | --- | --- | --- | --- |
| Australia | 24 | None | 26-28% reduction on 2005 levels | None at the Federal level |
| Australian Capital Territory | 97 | 100% | 2045 | Achieve zero emissions from the public health sector by 2040 |
| Tasmania | 96 | 100% | Achieved 2018 | None |
| South Australia | 52 | Removed | 2050* | None |
| Victoria | 24 | 50%* | 2050* | 5% of hospital electricity from on-site renewables by 2023; Work with Health Purchasing Victoria regarding state-wide power purchase agreements with renewables |
| Queensland | 14 | 50% | Aspirational 2050 | New health facilities shall *target* 20% of power sourced from alternative energy sources (*including fossil gas*) (2017) |
| New South Wales | 15 | None | Aspirational 2050 | All non-Local Health District's (LHD) facilities purchase 6% GreenPower; LHDs to complete at least one renewable energy installation/year *if* internal rate of return >12% can be achieved over life of project |
| Western Australia | 21 | None | Aspirational 2050 | None |
| Northern Territory | 8 | 50% | Aspirational 2050 | Assist government facilities (including health centres) to have more flexible financing options to enter into renewable energy projects |

\*Legislated targets.

## Discussion

Our study shows that less than 2.3% (14-94 GWh/yr) of Australian public hospital energy use was sourced from renewable energy between 2016/17 to 2018/19, with the vast majority arising from a Queensland Health power purchasing agreement (76% of total Australian public hospital renewable electricity) in 2018/19. In contrast, total energy consumption by public hospitals was considerable, consuming 4,122-4,132 GWh/yr over the same period. The equivalent energy usage would power approximately 630,000 average Australian homes each year.

The relatively small percentage uptake of renewables mean that public hospitals remain very large absolute emitters of greenhouse gases and harmful air particulates. Given public hospitals account for half of healthcare’s contribution to Australia’s total carbon footprint (3), then the contribution by public hospitals to Australia’s total greenhouse gas emissions remains considerable.

In comparison, England’s *NHS* accounts for about 5 percent of the UK’s total carbon footprint (8). More than 11 percent of the *NHS’s* 3,500 buildings (hospitals, health centres, GP surgeries and offices) are set to be powered by 100% renewable electricity in 2020 (23). In 2008, the UK established a national *Sustainable Development Unit* to coordinate and measure healthcare decarbonisation, developing a practical roadmap such that healthcare is a leading contributor toward achieving England’s 2050 net zero emissions targets.

At Australian state/territory level, renewable energy uptake by public hospitals has been piecemeal in contributing toward state/territory renewable and net zero ambitions. Together Victoria and New South Wales consumed 60% of healthcare direct energy, but each produce/purchase less than 1% renewable electricity. This is despite hospitals accounting for 26 percent of public energy use in Victoria (10) and over 50 percent of NSW Government emissions (9). In order for Victoria to achieve *at least 5 percent of hospital electricity to be sourced from on-site renewable energy by 2023* and contribute toward Victoria’s legislated 2050 net carbon zero target, large change will be required. Natural/Fossil gas consumption also presents a considerable challenge for Victorian public hospitals to wean with 48% of energy consumption attributed to natural/fossil gas use in 2018/19.

No change in renewable energy production/purchase by New South Wales hospitals between 2016/17 and 2018/19) may suggest that the NSW health sector target of *at least one new renewable energy installation per year if an internal rate of return of 12% or greater can be achieved* is impossible. Ultimately there is no requirement to use cleaner/renewable energy in New South Wales (24), beyond the expectation that hospitals have a role in evidence-based treatment, protection and promotion of health.

Queensland public hospitals were the largest purchaser/producer of renewable electricity (73.70 [9.47%] GWh, 2018/19). The whole-of-government approach increased purchasing power and facilitated the large-scale renewable purchase by public hospitals, although renewable energy is still more expensive according to state energy advisors (hospitals’ 24-hour operations demand both on- and off-peak electricity at increased cost). This approach has also been adopted by healthcare in the U.S with purchasing agreements enabling the construction of renewable energy farms and battery storage systems (7).

Queensland may demonstrate a pathway for other state/territory health systems who have not been able to overcome barriers to large-scale renewable purchasing. Victoria, for example, is seeking to achieve a major proportion of healthcare and state-wide carbon savings through large-scale renewables (10). However, GreenPower purchasing may not suit all states/territories. For example, the ACT, Tasmania and South Australia have already achieved high grid renewable electricity penetration, with ACT and South Australia reducing state-wide grid carbon intensity by more than 50% over the past decade through rapid uptake of renewables (18, 19). This may explain why relatively little renewable electricity has been produced/ purchased by public hospitals in these states/territories (range 0.42 - 1.2%, 2018/19).

However, large amounts of fossil/natural gas are still consumed nationally, inclusive of states/territories with cleaner electricity supply such as South Australia and the ACT. In this instance alternative pathways forward are being demonstrated by healthcare institutions in the U.S, providing strong business cases for hospital carbon neutrality, whilst expanding operations (7).

Other economic sectors in Australia, such as tertiary education, have set 100% renewable electricity targets (25). Renewables investment by individual tertiary institutions is occurring at a scale equal or larger than entire state/territory public hospital efforts. Compared to all Victorian public hospitals (11 GWh, 2018/19), Monash University is alone purchasing/producing five times the amount of renewable electricity each year (55 GWh of GreenPower; 3.8-6.3 GWh of solar PV, 2015/16 to 2018/19). Other universities (Swinburne University 90.4 GWH/yr; University of New South Wales 124 GWh/yr) have entered long-term renewable Power Purchase Agreements that eclipse the efforts of the entire Australian public hospital system.

Obtaining the information for this study was not easy, and therefore our results are subject to several assumptions and limitations. We calculated the best-case scenario for the percentage of renewable energy produced/purchased, often based on peak-solar output. Underreporting of total hospital energy data was possible in Western Australia, Northern Territory, Australian Capital Territory and Queensland with very small rural/regional facilities not included in the supplied data. Centralised management systems of public hospital energy are not established in some jurisdictions. As a consequence, raw data provided by state energy analysts often contained multiple estimates. We did not consider indirect hospital energy consumption, such as procurement of services, equipment and consumables not directly controlled or owned by the hospital. We also did not examine private hospital energy use due to commercial in confidence. Future analyses should consider data collection methodology in the absence of robust state-wide energy management systems and broaden the defined scope of energy consumption.

Despite these limitations, our study is original in seeking to understand how rapidly Australian hospitals are moving towards renewable energy as a source of electricity supply and to compare the performance of the health sector against international leaders and other economic sectors. This provides an important baseline for leaders within Australian healthcare to consider the sectors carbon footprint and the extent to which public hospitals are contributing to achieving net zero emissions targets or aligning energy choices with the remit of the health sector.

The Australian healthcare sector is being left behind by global healthcare leaders and other economic sectors in the uptake of renewable energy. The Hippocratic oath ‘*First Do No Harm*’ is central to the day-to-day practice of medicine. Climate change has been identified as the major health threat of this century. Therefore, in order to decouple hospital energy from the health and health system impacts of climate change, hospitals must address their energy choices. Queensland has demonstrated what can occur, through purchasing large-scale renewable energy to power its public hospitals. This study supports improved leadership by healthcare and encourages doctors and other healthcare workers to encourage their energy procurers to follow suit. Ultimately a high-quality sustainable healthcare system could be achieved through developing a roadmap toward reducing healthcare CO2e emissions that meets the needs of today, without compromising the needs of tomorrow.

## Data Availability

The data that support the findings of this study are available from the corresponding author, HB, upon reasonable request.

## List of abbreviations

HVAC: Heating, ventilation and air condition
NHS: National Health Service
NABERS: National Built Energy Rating System
NGER: National Greenhouse & Energy Reporting
GWh: gigawatt-hours
GJ: gigajoules
LPG: Liquid propane gas
LHD: Local health district
COAG: Council of Australian Governments

## References

1. Costello A, Abbas M, Allen A, Ball S, Bell S, Bellamy R, et al. Managing the health effects of climate change. The Lancet. 2009;373(9676):1693–733.

2. Watts N, Adger WN, Agnolucci P, Blackstock J, Byass P, Cai W, et al. Health and climate change: policy responses to protect public health. The Lancet. 2015;386(10006):1861–914.

3. Malik A, Lenzen M, McAlister S, McGain F. The carbon footprint of Australian health care. The Lancet Planetary Health. 2018;2(1):e27–e35.

4. Department of Climate Change and Energy Efficiency. Baseline Energy Consumption and Greenhouse Gas Emissions - Part 1. Canberra: Commonwealth of Australia; 2012.

5. Department of Health & Human Services. Energy use in Victorian public healthcare services: State Government of Victoria; 2018 [Available from: https://www2.health.vic.gov.au/hospitals-and-health-services/planning-infrastructure/sustainability/energy/energy-use.

6. Karliner J, Slotterback S, Boyd R, Ashby B, Steele K. Health Care’s Climate Footprint: How the Health Sector Contributes to the Global Climate Crisis and Opportunities for Action. https://noharm-uscanada.org/ClimateFootprintReport: Health Care Without Harm; 2019.

7. Chen A, Murthy V. How Health Systems Are Meeting the Challenge of Climate Change. Harvard Business Review. 2019.

8. Sustainable Development Unit (SDU). Carbon print update for NHS in England 2015. Cambridge, UK: SDU; 2016.

9. NSW Auditor General. Building energy use in NSW public hospitals. Sydney: Audit Office of NSW; 2013.

10. Victorian Health and Human Services Building Authority. Environmental Sustainability Strategy: 2018-19 to 2022-23. State of Victoria: Victorian State Government; 2018.

11. NABERS. Annual Report 2018/19. National Australian Built Environment Rating System,; 2019.

12. Clean Energy Regulator. National Greenhouse and Energy Reporting (NGER) reporting thresholds: Australian Government; 2018 [Available from: http://www.cleanenergyregulator.gov.au/NGER/Reporting-cycle/Assess-your-obligations/Reporting-thresholds.

13. AIHW. Hospital resources 2017–18: Australian hospital statistics. Australian Institute of Health and Welfare, Australian Government; 2019.

14. Department of Environment and Energy. National Greenhouse Accounts Factors. https://www.environment.gov.au/system/files/resources/5a169bfb-f417-4b00-9b70-6ba328ea8671/files/national-greenhouse-accounts-factors-july-2017.pdf:CommonwealthofAustralia;2017.

15. Solar Choice. How much energy will my solar cells produce? 2019 [Available from: https://www.solarchoice.net.au/blog/how-much-energy-will-my-solar-cells-produce/.

16. State of NSW, Office of Environment and Heritage. Cogeneration feasibility guide. Sydney: NSW Government; 2014.

17. IPCC. Summary for Policymakers of IPCC Special Report on Global Warming of 1.5°C approved by governments. Online: United Nations; 2018. Contract No.: 4.

18. ACT Government. ACT Climate Change Strategy 2019–25. Canberra, Australian Capital Territory; 2019.

19. Clean Energy Council. Clean Energy Australia Report. 2020.

20. Northern Territory Government. Northern Territory Climate Change Response: Towards 2050. Darwin, Northern Territory; 2019.

21. NSW Government. NSW Government Resource Efficiency Policy. Office of Environment and Heritage; 2019.

22. Queensland Health. Capital Infrastructure Requirements. Queensland Government; 2017.

23. NHS Property Services. Over 10 per cent of NHS estate to switch to renewable energy Hospital Times 2020 [Available from: https://www.hospitaltimes.co.uk/over-10-per-cent-of-nhs-estate-to-switch-to-renewable-energy/.

24. Building Energy Use in NSW Public Hospitals [press release]. https://www.audit.nsw.gov.au/media-release/building-energy-use-in-nsw-public-hospitals2013.

25. Clean Energy Council. Renewable energy powering Australian education 2019 [Available from: https://www.cleanenergycouncil.org.au/news/renewable-energy-powering-australian-education.

